# Seroprevalence Convergence Does Not Reflect Transmission Equity: Persistent Socioeconomic Disparities in COVID-19 Force of Infection in Canada

**DOI:** 10.64898/2026.01.02.26343363

**Authors:** Abdullah Hassan, Emmanuel Issa Nassrallah, David N. Fisman

**Author notes:** Address reprint requests and correspondence to: David Fisman, MD MPH FRCP(C), Room 686, 155 College Street, Toronto, Ontario, M5T 3M7. Both authors contributed equally to the work (Abdullah Hassan, Emmanuel Issa Nassrallah). **Code Availability:** Analysis code is available at https://github.com/fismanda/covid19-seroprev-equity and permanently archived at https://doi.org/10.5281/zenodo.18064548. **Data Availability:** Seroprevalence data used in this study are publicly available from the COVID-19 Immunity Task Force (CITF) Data Portal at https://portal.citf.mcgill.ca.

## Abstract

**Background:** Socioeconomic disparities in COVID-19 outcomes have been widely documented, but evidence regarding inequities in SARS-CoV-2 transmission remains mixed. In Canada, infection-induced seroprevalence appeared to converge across socioeconomic strata by late 2022, raising questions about whether transmission inequities diminished during the Omicron period.

**Aim:** To assess whether apparent convergence in SARS-CoV-2 seroprevalence reflects true equity in transmission or masks persistent socioeconomic disparities in force of infection.

**Methods:** We analysed serial cross-sectional SARS-CoV-2 seroprevalence data (anti-nucleocapsid antibodies) from Canadian Blood Services donors collected between April 2021 and April 2023 and stratified by area-level material deprivation quintile. We fitted a dynamic susceptible–infected model with sero-reversion to the full seroprevalence time series, estimating quintile-specific forces of infection before and after the emergence of the Omicron variant (January 2022). Models allowing differential Omicron-related amplification by socioeconomic status were compared using likelihood-based criteria.

**Results:** During the pre-Omicron period, force of infection increased monotonically with material deprivation; the most deprived quintile experienced a 71% higher force of infection than the least deprived (incidence rate ratio (IRR): 1.71; 95% CI: 1.60–1.83). Following Omicron emergence, force of infection increased markedly in all quintiles but by differing magnitudes. Relative increases were largest in the least deprived quintile (48.5-fold) and smallest in the most deprived quintile (31.8-fold), resulting in compression of the socioeconomic gradient (Q5 vs Q1 IRR: 1.12; 95% CI: 1.11–1.14). Despite this compression, materially deprived populations continued to experience elevated transmission risk.

**Conclusion:** Convergence in SARS-CoV-2 seroprevalence across socioeconomic strata masked persistent inequalities in force of infection. Dynamic modelling demonstrates that apparent equity arose from differential amplification of transmission during the Omicron period rather than from elimination of underlying socioeconomic disparities.

## INTRODUCTION

The COVID-19 pandemic disproportionately affected marginalized populations worldwide, with well-documented gradients in infection risk, severe outcomes, and mortality by socioeconomic status (SES) ^1–4^. In Canada, as in many high-income countries, lower SES was consistently associated with higher rates of COVID-19 hospitalization and death throughout the pandemic.^6–8,5–7^. These disparities reflect structural determinants of health including occupational exposure risk (essential workers unable to work remotely), household crowding, reduced access to healthcare, and barriers to implementing protective measures such as physical distancing ^1,5^.

Serological surveillance offers unique insights into population-level infection dynamics by measuring antibody prevalence, which captures both diagnosed and undiagnosed infections ^4,8–11^. Unlike case-based surveillance, which is subject to differential testing access and health-seeking behavior ^12^, seroprevalence reflects cumulative infection exposure across populations. The COVID-19 Immunity Task Force (CITF) conducted serial serosurveys of Canadian blood donors from 2020 through 2023, providing detailed temporal data on infection-induced (anti-nucleocapsid) antibody prevalence stratified by material deprivation quintiles ^4,8^. These data are publicly available ^13^.

Previous analyses of these data by O’Brien and colleagues ^8^, documented significant SES gradients in COVID-19 seroprevalence during the pre-Omicron period (2020-2021), with higher infection-induced antibody prevalence in materially deprived areas. However, by late 2022 and early 2023, seroprevalence estimates appeared to converge across SES strata, reaching 75-85% in all quintiles ^8^. This convergence might suggest that socioeconomic disparities in COVID-19 transmission diminished as the pandemic progressed, particularly following the emergence of the highly transmissible Omicron variant in late 2021.

However, seroprevalence convergence does not necessarily imply epidemiological equity. When examining cumulative measures like seroprevalence, populations with higher baseline transmission rates will reach saturation earlier, potentially masking persistent disparities in the underlying force of infection (the instantaneous per capita rate at which susceptible individuals acquire infection)²⁰⁻²². As susceptible pools become depleted in high-transmission populations, rates of seroprevalence change may slow even while the force of infection remains elevated. This phenomenon has been observed in other infectious disease contexts^14,15^ but has not been formally evaluated for COVID-19 socioeconomic gradients.

Moreover, the Omicron variant’s emergence fundamentally altered pandemic dynamics. With several-fold increase in reproduction number compared to Delta^16^, combined with substantial immune evasion^17^, Omicron generated widespread community transmission even in highly vaccinated populations. The extent to which this universal transmission pressure affected different socioeconomic groups, whether it amplified, maintained, or compressed existing disparities, remains unclear.

To address these questions, we employed a dynamic compartmental modeling approach to estimate baseline force of infection and its amplification during the Omicron era across SES quintiles. Rather than comparing static seroprevalence estimates, we fit a two-phase susceptible-infected model to the complete time series of seroprevalence data, accounting for susceptible depletion and potential sero-reversion. This approach allows us to disentangle baseline transmission disparities from the universal effects of increased variant transmissibility, and to determine whether observed seroprevalence convergence reflects true epidemiological equity or mathematical artifact.

We sought to answer two primary questions. First, did baseline force of infection differ by socioeconomic status throughout the COVID-19 pandemic in Canada? Secondly, we asked how the emergence of Omicron SARS-CoV-2 variants affected transmission across socioeconomic strata.

## METHODS

### Study Design and Data Sources

We conducted a dynamic modeling analysis of serial cross-sectional seroprevalence data from the Canadian Blood Services (CBS) COVID-19 serosurveillance program. CBS conducted repeated serosurveys of blood donors across Canada from June 2020 through April 2023, with samples tested for antibodies to the SARS-CoV-2 nucleocapsid protein (anti-N), which indicates prior infection. These data were made publicly available through the COVID-19 Immunity Task Force (CITF) and have been previously described ^4,8,13^.

Donor postal codes were deterministically linked to area-level material deprivation indices from the Canadian Index of Multiple Deprivation ^4,18^, categorized into quintiles (Q1 = least deprived to Q5 = most deprived). We restricted our analysis to CBS data only to ensure consistency in sampling methodology, laboratory assays, and participant characteristics throughout the study period. All serosurveys assessing infection-induced seroprevalence (anti-N antibodies) stratified by material deprivation quintile were included.

### Study Population and Period

The study population consisted of adult blood donors (≥18 years) who met Canadian Blood Services eligibility criteria. Blood donors represent a healthier subset of the general population and findings should be interpreted as representative of this specific population rather than all Canadians. Although community transmission of SARS-CoV-2 began in March 2020, and some seroprevalence data were available from the early pandemic, serological data including all 5 material deprivation quintiles spanned April 2021 through April 2023, encompassing the mid-pandemic period through to the mid-Omicron era. We defined two pandemic periods based on variant epidemiology: pre-Omicron (March 2020 - December 31, 2021), during which wild-type, Alpha, and Delta variants circulated; and Omicron (January 1, 2022 - April 2023), following the emergence of the Omicron variant in late December 2021. This binary classification reflects the substantial shift in transmission dynamics associated with Omicron’s increased transmissibility and immune evasion capabilities.

### Mathematical Model

We developed a two-compartment susceptible-infected (SI) model with sero-reversion to estimate force of infection from observed seroprevalence trajectories. This framework does not treat seropositivity as permanent, but rather, accounts for antibody waning and the fact that seroprevalence represents a dynamic balance between seroconversion and sero-reversion.

The model is defined by the following difference equations:

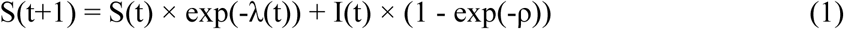

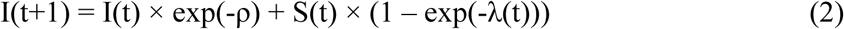

where S(t) represents the proportion susceptible (seronegative) at time t, I(t) represents the proportion infected (seropositive), λ(t) is the time-varying force of infection (per capita rate of infection acquisition), and ρ is the sero-reversion rate (per capita rate of antibody loss).

We initialized the model at March 1, 2020 (S(0) = 1, I(0) = 0), corresponding to the start of community transmission in Canada. The model simulates seroprevalence dynamics forward from this baseline through April 2023. As seroprevalence data stratified by material deprivation quintile were not available for all quintiles until April 2021, parameter estimation was performed using data from April 21, 2021 onwards, when all five quintiles had available observations. This approach ensures the model accounts for the full infection history while fitting to the period with complete quintile-specific data. Model fitting is described in detail in the **Supplementary Appendix 1**.

Because published estimates of anti-nucleocapsid (anti-N) antibody waning vary widely ^19–24^, we conducted sensitivity analyses to assess whether conclusions depended on assumptions about sero-reversion. In addition to estimating the sero-reversion rate (ρ) directly from the data, we refit the saturated model with ρ fixed to literature-based values spanning the reported range of anti-N durability (half-lives of approximately 2 to 8 months). For each scenario, we re-estimated quintile-specific forces of infection and compared socioeconomic gradients across pre-Omicron and Omicron periods. Full details are provided in **Supplementary Appendix 2**.

All analyses were conducted using R version 4.3.0 (R Foundation for Statistical Computing, Vienna, Austria). Maximum likelihood estimation used the bbmle package, data manipulation used dplyr and tidyr, and visualizations used ggplot2. Code is available at https://github.com/fismanda/covid19-seroprev-equity and archived at https://doi.org/10.5281/zenodo.18064548.

### Ethical Considerations

This study analyzed de-identified, aggregated seroprevalence data that were publicly released by the COVID-19 Immunity Task Force. No individual-level data were accessed. The original serosurveillance program received ethics approval from Canadian Blood Services and participating institutions; details have been published previously ^4,8^.

## RESULTS

### Study Sample and Data Characteristics

The analysis included 410 serosurvey observations from Canadian Blood Services spanning April 21, 2021 to April 26, 2023 (when all five material deprivation quintiles had available data). A total of 82 measurements with complete quintile data were available during this time period.

Sample sizes per observation ranged from approximately 1,500 to 20,000 blood donors, with larger samples in less deprived quintiles reflecting the demographic composition of blood donors. Crude seroprevalence estimates increased substantially over time in all quintiles, rising from approximately 5% in April 2021 to 75-85% by April 2023. Visual inspection of temporal trajectories suggested converging seroprevalence across socioeconomic strata by late 2022, consistent with widespread Omicron transmission (**Figure 1**). We compared fits of the parsimonious model (which constrained all quintiles to experience the same proportional increase during Omicron) and a saturated model in which Omicron’s impact varied was allowed to vary by socioeconomic status and identified strong improvements in fit with the saturated model, with difference in Akaike Information Criterion (AIC) between the two models of - 194.17. Model fitting results are described further in the **Supplementary Appendix 1**.

**Figure 1.**
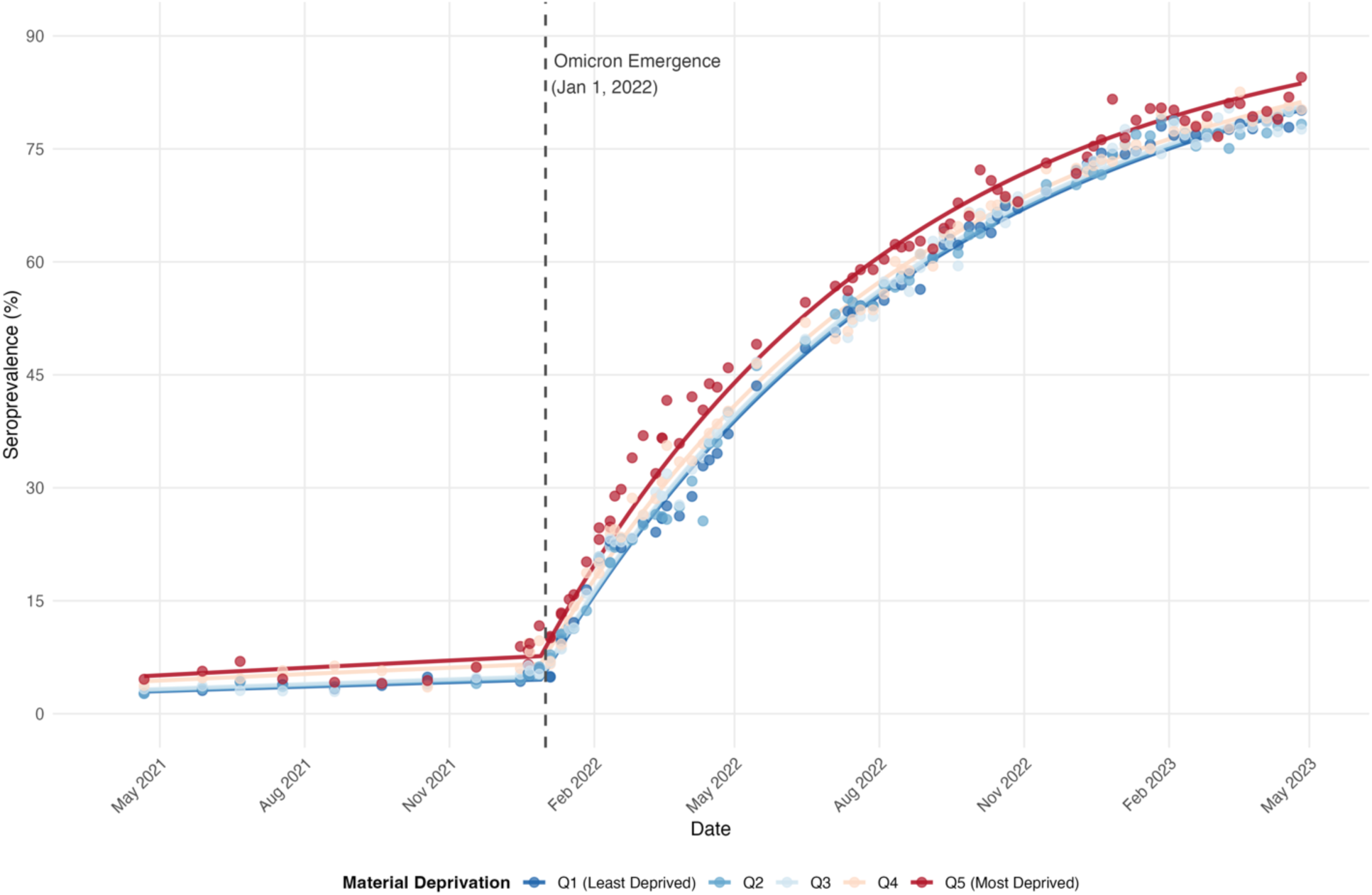
Infection-induced SARS-CoV-2 seroprevalence by material deprivation quintile, Canada, 2021–2023.

For the saturated model, the estimated sero-reversion rate was ρ = 0.00193 per week (95% CI: 0.00171, 0.00217), corresponding to an antibody half-life of approximately 83 months (6.9 years; 95% CI: 74-93 months). A similarly low rate of sero-reversion was estimated in the parsimonious model (ρ = 0.00193 per week).

During the pre-Omicron period, force of infection demonstrated a clear socioeconomic gradient, with higher transmission rates in more materially deprived areas (**Table 1** and **Figure 2**). The baseline force of infection in Q1 (least deprived) was 0.00056 per week, corresponding to approximately 0.056% of the susceptible population becoming infected per week. The second-least deprived quintile (Q2) did not differ significantly from Q1 (IRR 1.04, 95% CI 0.98-1.11). However, more deprived quintiles experienced progressively greater force of infection: Q3 IRR 1.09 (95% CI 1.02-1.17); Q4 IRR 1.48 (95% CI 1.38-1.58); Q5 (most deprived) IRR 1.71 (95% CI 1.60-1.83). A meta-regression model affirmed a log-linear dose-response relationship between increasing deprivation and increasing force of infection (relative IRR per quintile 1.204, 95% CI 1.014-1.429, P = 0.043). The most deprived quintile (Q5) experienced 71% higher force of infection than the least deprived quintile during the pre-Omicron period, representing substantial socioeconomic inequality in transmission risk.

**Figure 2.**
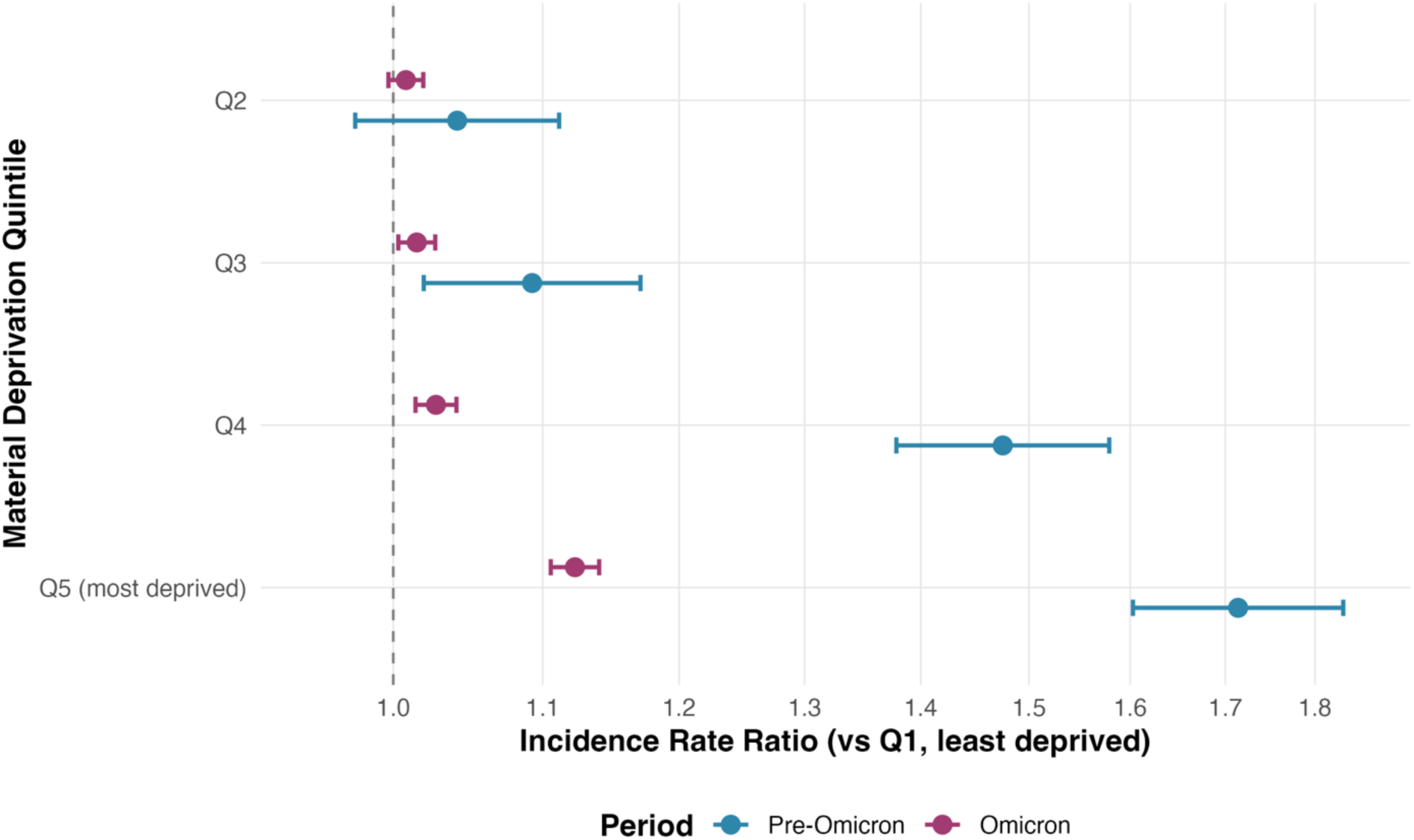
Socioeconomic gradient in SARS-CoV-2 force of infection before and during the Omicron period.

**Table 1.**
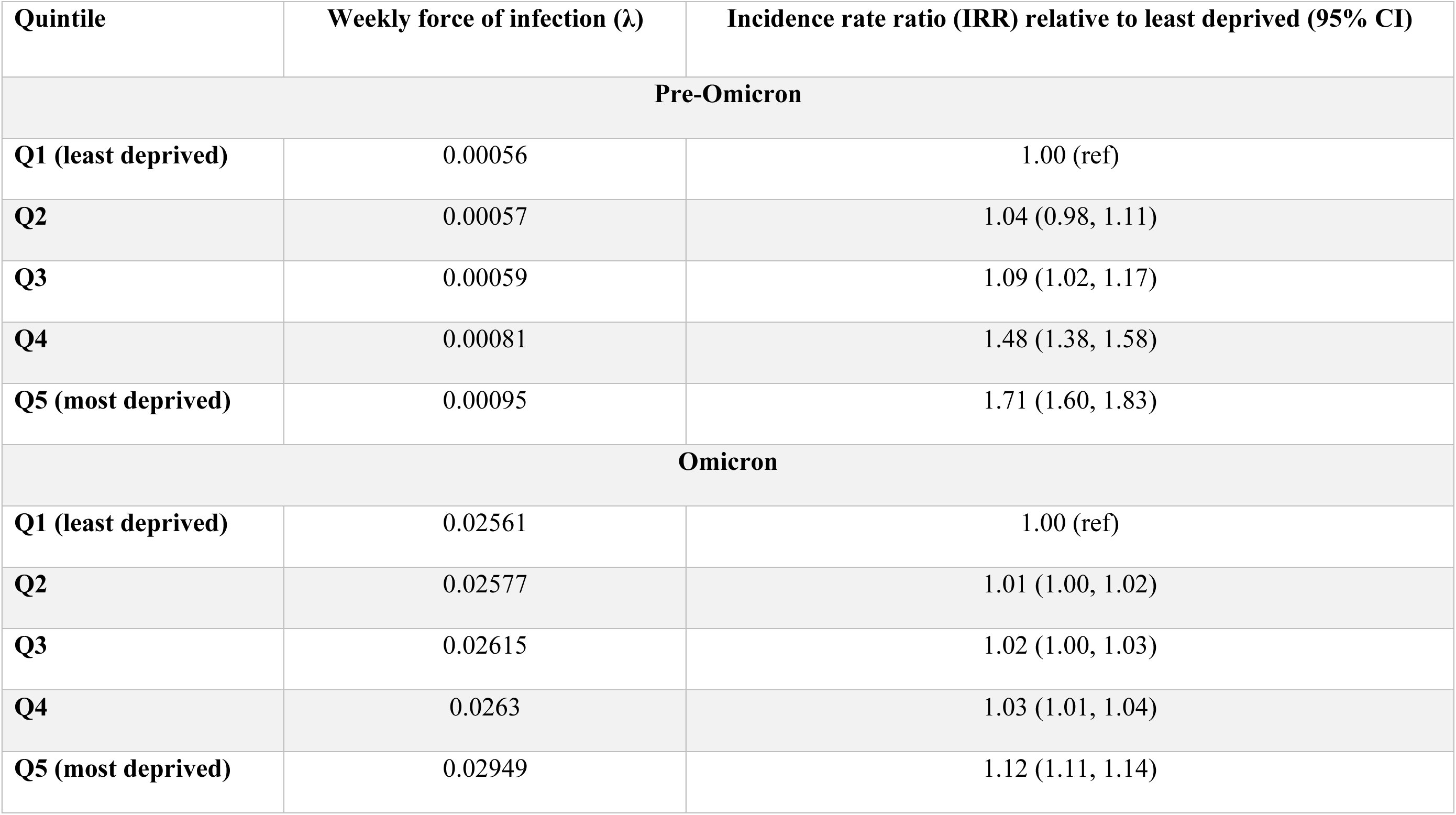
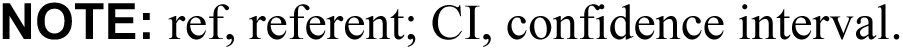
Force of infection estimates from saturated model, and incidence rate ratios for infection relative to least deprived quintile, during pre-Omicron and Omicron periods in Canada.

| Quintile | Weekly force of infection ($\lambda$) | Incidence rate ratio (IRR) relative to least deprived (95% CI) |
| --- | --- | --- |
| **Pre-Omicron** |  |  |
| **Q1 (least deprived)** | 0.00056 | 1.00 (ref) |
| **Q2** | 0.00057 | 1.04 (0.98, 1.11) |
| **Q3** | 0.00059 | 1.09 (1.02, 1.17) |
| **Q4** | 0.00081 | 1.48 (1.38, 1.58) |
| **Q5 (most deprived)** | 0.00095 | 1.71 (1.60, 1.83) |
| **Omicron** |  |  |
| **Q1 (least deprived)** | 0.02561 | 1.00 (ref) |
| **Q2** | 0.02577 | 1.01 (1.00, 1.02) |
| **Q3** | 0.02615 | 1.02 (1.00, 1.03) |
| **Q4** | 0.0263 | 1.03 (1.01, 1.04) |
| **Q5 (most deprived)** | 0.02949 | 1.12 (1.11, 1.14) |

**NOTE:** ref, referent; CI, confidence interval.

Following Omicron emergence in January 2022, force of infection increased dramatically in all socioeconomic strata, but the magnitude of increase varied substantially by material deprivation level (**Table 2** and **Figure 3**). The socioeconomic gradient became markedly less steep, though persistent disparities remained. During the Omicron period, Q2 did not differ significantly from Q1 (IRR 1.01, 95% CI 1.00-1.02), while more deprived quintiles showed progressively elevated risk: Q3 IRR 1.02 (95% CI 1.00-1.03); Q4 IRR 1.03 (95% CI 1.01-1.04); Q5 (most deprived) IRR 1.12 (95% CI 1.11-1.14). Meta-regression no longer revealed a significant log-linear relationship between quintile and force of infection (relative IRR per quintile 1.042, 95% CI 0.965-1.125, P = 0.148).

**Figure 3.**
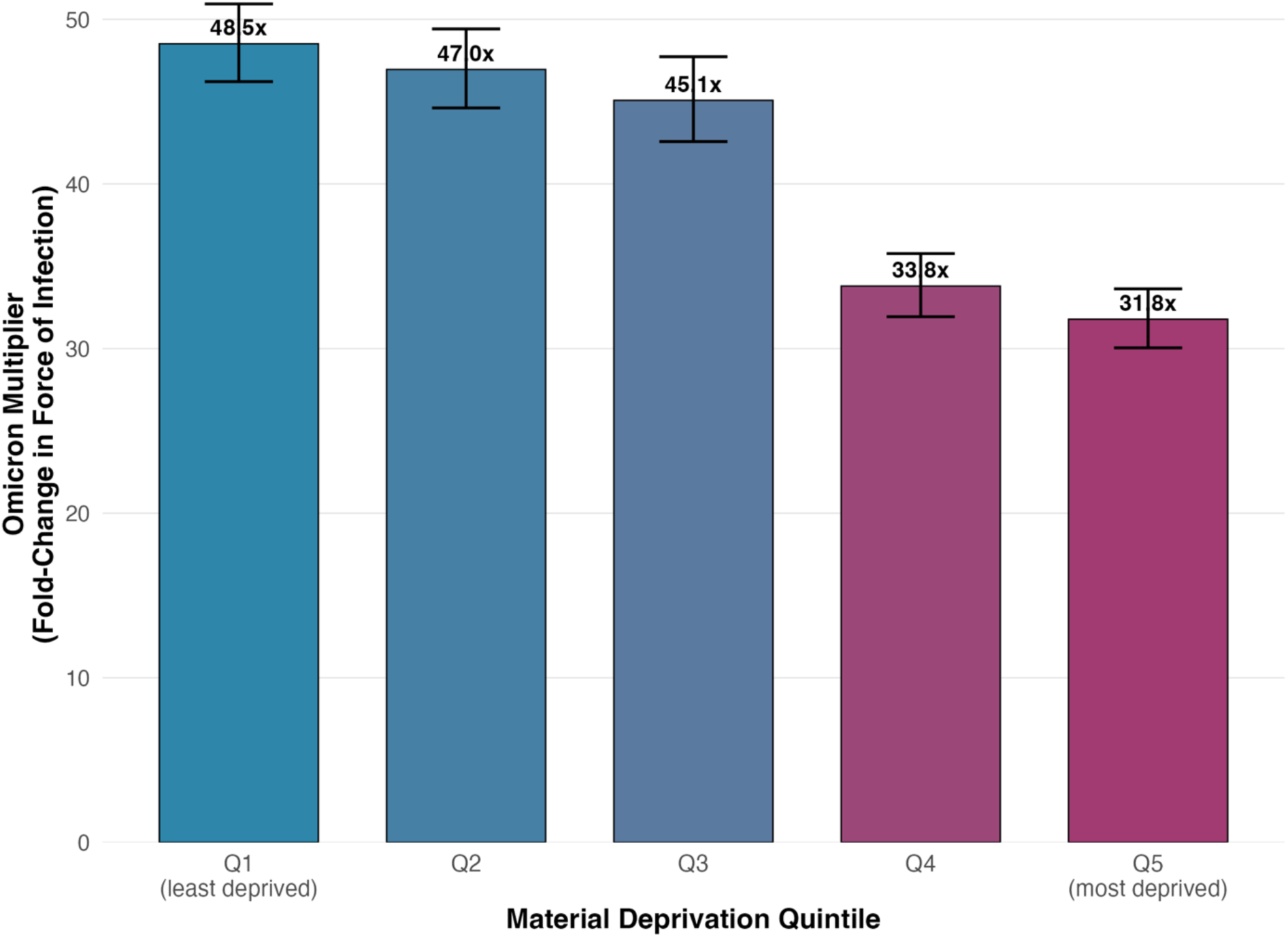
Differential amplification of SARS-CoV-2 transmission following Omicron emergence by material deprivation quintile.

**Table 2.** Comparison of force of infection and incidence rate ratios relative to least deprived quintile with Omicron emergence.

| **Quintile** | **Absolute increase in weekly $\lambda$ with Omicron** | **Omicron multiplier (95% CI)** | **Ratio of ratios\* (95% CI)** | **P-value for ratio of ratios** |
| --- | --- | --- | --- | --- |
| **Q1 (least deprived)** | 0.02505 | 48.52 (47.23, 49.84) | 1.00 (ref) | — |
| **Q2** | 0.0252 | 46.95 (45.71, 48.22) | 0.97 (0.90, 1.04) | 0.36 |
| **Q3** | 0.02557 | 45.07 (43.85, 46.31) | 0.93 (0.86, 1.00) | 0.053 |
| **Q4** | 0.02549 | 33.80 (33.02, 34.59) | 0.70 (0.65, 0.75) | <0.001 |
| **Q5 (most deprived)** | 0.02854 | 31.78 (31.04, 32.53) | 0.66 (0.61, 0.71) | <0.001 |

**NOTE:** ref, referent; CI, confidence interval; $\lambda$ , force of infection.
\*Ratio of quintile-specific incidence rate ratio (IRR) after Omicron emergence to IRR before Omicron.

The Q5-to-Q1 incidence rate ratio decreased from 1.71 pre-Omicron to 1.12 during Omicron, representing a 35% relative compression of the socioeconomic gradient. To formally test whether this compression was statistically significant, we calculated IRR ratios (Omicron IRR / Pre-Omicron IRR) for each quintile. These ratios revealed significant gradient compression for the most deprived quintiles: Q2 ratio 0.97 (95% CI 0.90-1.04, P = 0.360); Q3 ratio 0.93 (95% CI 0.86-1.00, P = 0.053); Q4 ratio 0.70 (95% CI 0.65-0.75, P < 0.001); Q5 ratio 0.66 (95% CI 0.61-0.71, P < 0.001). Critically, despite this compression, the most deprived quintile maintained 12% higher force of infection than the least deprived during Omicron, demonstrating that apparent seroprevalence convergence masked persistent socioeconomic disparities in ongoing transmission.

The mechanism underlying gradient compression was differential amplification: quintile-specific increases in force of infection varied inversely with baseline transmission levels. Omicron multipliers, calculated as the ratio of Omicron-era to pre-Omicron force of infection, revealed striking heterogeneity (**Table 2**). The least deprived quintile experienced a 48.52-fold increase (95% CI 47.23-49.84), while the most deprived quintile experienced a 31.78-fold increase (95% CI 31.04-32.53). This 53% difference in amplification effects (Q1/Q5 multiplier ratio = 1.53) explains both the dramatic improvement in model fit for the saturated specification and the observed seroprevalence convergence. The differential amplification represents “leveling down” with affluent populations experienced larger proportional increases to approach (but not reach) transmission levels already experienced by disadvantaged populations, rather than genuine reduction in health inequities.

Sensitivity analyses demonstrated that the magnitude and direction of socioeconomic gradients in force of infection were robust across a 40-fold range of assumed sero-reversion rates. Although models constrained to faster antibody waning fit the data substantially worse, estimates of pre-Omicron and Omicron-era inequality, Omicron-era gradient compression, and differential amplification were qualitatively unchanged (**Supplementary Appendix 2**).

## DISCUSSION

Using dynamic modeling of serial seroprevalence data from Canadian blood donors, we found that socioeconomic gradients in COVID-19 force of infection persisted throughout the pandemic but compressed substantially following Omicron emergence. During the pre-Omicron period (2020-2021), the most materially deprived quintile experienced 71% higher force of infection compared to the least deprived quintile (IRR = 1.71). This gradient narrowed to 12% during the Omicron era (2022-2023; IRR = 1.12), not through equity improvements but via differential amplification: the least deprived populations experienced a 49-fold increase in force of infection while the most deprived experienced a 32-fold increase.

Critically, this analysis demonstrates that seroprevalence convergence, the observation that cumulative infection levels became similar across socioeconomic strata by late 2022, masks persistent inequalities in underlying transmission dynamics. While crude seroprevalence estimates suggested diminishing disparities, force of infection modeling revealed that materially deprived populations maintained elevated transmission risk even at high levels of population immunity.

Understanding whether seroprevalence convergence reflects genuine equity or mathematical artifact has critical implications for pandemic preparedness and health policy. If convergence truly reflected diminished disparities, it would suggest that structural interventions, variant characteristics, or population saturation successfully mitigated socioeconomic gradients in transmission. Conversely, if convergence masks persistent disparities in force of infection, it indicates that inequities remained throughout the pandemic but became mathematically obscured as population seropositivity approached saturation.

Our findings demonstrate the latter. Seroprevalence convergence arose from differential depletion of seronegative populations rather than elimination of transmission disparities. Because the most deprived quintile experienced higher cumulative infection during the pre-Omicron period, a larger proportion of this population was already seropositive when Omicron emerged.

Consequently, even with 12% higher ongoing force of infection throughout the Omicron period, the rate of increase in seroprevalence appeared similar across quintiles, creating an illusion of equity in cross-sectional or static longitudinal comparisons. As populations approach saturation (75-85% seroprevalence in the CBS dataset), rates of seroprevalence change necessarily slow regardless of ongoing transmission intensity due to an ever-smaller seronegative denominator. A population experiencing higher force of infection will reach saturation earlier, at which point its seroprevalence trajectory will plateau even as transmission remains elevated. This creates a fundamental measurement problem: the very populations that experienced highest transmission early in the pandemic appear to achieve equity precisely because they were disproportionately affected.

A potential concern is that the sero-reversion rate estimated in our primary analysis implies substantially longer persistence of anti-nucleocapsid antibodies than reported in many individual-level cohort studies. However, extensive sensitivity analyses demonstrate that our substantive conclusions do not depend on this parameter choice. Across a 40-fold range of sero-reversion rates drawn from pre-Omicron and Omicron-era literature, we consistently observed strong pre-Omicron socioeconomic gradients in force of infection, persistent but compressed disparities during Omicron, and differential amplification of transmission among less deprived populations. While faster waning assumptions degraded overall model fit, estimates of relative transmission risk were remarkably stable, reflecting the fact that seroprevalence trajectories during this period were dominated by high force of infection rather than antibody loss. These findings indicate that apparent seroprevalence convergence cannot be explained by modeling artifacts related to antibody kinetics and instead reflects genuine—but obscured—inequalities in transmission dynamics.

Our findings both complement and extend prior analyses of the CITF dataset. O’Brien and colleagues, documented significant socioeconomic gradients in COVID-19 seroprevalence during 2020-2021, with higher infection-induced antibody prevalence in materially deprived areas^8^. They observed apparent convergence by late 2022, raising questions about whether pandemic inequities diminished over time. However, the gradients in force of infection that we report here align with socioeconomic patterns in COVID-19 mortality in Canada. Analysis of linked Census-mortality data demonstrated that low-income status substantially increased COVID-19 mortality risk, with particularly pronounced effects among racialized Canadians, and Indigenous peoples. Among Black Canadians, low-income residence was associated with 3-fold higher mortality compared to higher-income Black Canadians, and individuals who were both Black and impoverished experienced a 5-fold elevated risk of mortality relative to those who were neither. First Nations people experienced 4.5-fold higher COVID-19 mortality than non-Indigenous people, with intersecting vulnerabilities including unsuitable housing and low income further elevating risk.

The CBS data are also consistent with our earlier observation that Canada’s pandemic response during the pre-Omicron period was relatively successful compared to peer nations^25^, notwithstanding the persistent socioeconomic gradients we document here. The remarkable surge in infection seroprevalence with Omicron variant emergence (a 49-fold increase in force of infection among the least deprived quintile and a 32-fold increase in the most deprived quintile) likely reflects the confluence of Omicron-related immune evasion, increased infectivity, and elimination of community-level SARS-CoV-2 control measures (including masking and vaccination requirements and school closures and work-from-home provisions). It is worth noting that the dramatic difference in force of infection between least and most deprived quintiles prior to Omicron emergence may serve as indirect evidence of the effectiveness of such structural advantages as remote work and private transportation enjoyed by more affluent Canadians early in the pandemic for the prevention of infection.

The socioeconomic gradients described here are not unique to Canada, nor to the COVID-19 pandemic. Franke’s analysis of the 1918 influenza pandemic in the German Kingdom of Wurttemberg demonstrated that low-income parishes experienced significantly higher mortality increases than high-income parishes, with middle- and high-income parishes recording substantially lower excess mortality during the pandemic year^26^.

Similarly, Mamelund’s detailed study of the 1918 pandemic in Bergen, Norway documented striking temporal variation in morbidity by socioeconomic status across pandemic waves.Low-income residents (classified based on apartment size as a proxy for crowding and economic status) experienced higher morbidity during the first wave in summer 1918, while high-income residents experienced higher morbidity during the second wave in fall 1918. Mamelund attributed this crossover to differential prior immunity: those with higher first-wave exposure gained protection against subsequent waves ^27^.

However, these historical analyses, like more recent COVID-19 seroprevalence studies, measured cumulative infection or crude morbidity rates rather than force of infection among remaining susceptibles. Even Mamelund’s observed “crossover,” where high-income groups appeared to surpass low-income groups in second-wave morbidity, could reflect the same mathematical artifact we document for COVID-19: if low-income residents maintained higher force of infection in the second wave, but a larger proportion were already immune from first-wave exposure, crude morbidity rates would appear similar or even reversed while underlying transmission risk remained elevated for susceptible individuals within disadvantaged groups. Our findings suggest that apparent convergence, or even crossover, in aggregate measures like hospitalization or mortality may obscure persistent disparities in underlying transmission risk that can only be detected through dynamic modeling of force of infection.

Several important limitations to this work warrant consideration. First, our analysis was restricted to Canadian blood donors, who represent a healthier, voluntary subset of the population and likely underrepresent individuals experiencing severe marginalization. The socioeconomic gradients we observed may underestimate true population-level disparities. Second, socioeconomic status was measured at the area level rather than individually, introducing ecological bias that may attenuate true associations. Third, we could not adjust for potential confounders including age, occupation, household composition, or race/ethnicity due to the aggregated nature of seroprevalence data. Finally, our binary classification of pre-Omicron versus Omicron periods simplifies a more complex variant landscape that included N501Y mutant (Alpha, Beta, Gamma), and Delta waves of SARS-CoV-2. Finer temporal resolution might reveal additional heterogeneities, though data sparsity in earlier waves would limit statistical power.

Future research should extend this analytical approach to other populations and settings to assess generalizability, and whether similar patterns of gradient compression through differential amplification occurred in other countries. Additionally, linking force of infection estimates to severe outcome data would clarify how transmission disparities translate to clinical burden. Finally, understanding mechanisms behind differential Omicron amplification requires integrating transmission modeling with detailed behavioral and environmental data on ventilation, mask quality, workplace exposures, and household crowding to inform targeted interventions for future respiratory pathogen threats.

In summary, we find that socioeconomic gradients in COVID-19 force of infection persisted throughout the pandemic in Canada, albeit with compression during the Omicron phase of the pandemic that reflected differential amplification: less deprived populations experienced larger proportional increases in transmission as variant transmissibility overwhelmed previously effective protections and public health control measures were discontinued. This compression occurred through worsening outcomes for affluent populations rather than improved equity, representing a “leveling down” phenomenon. Critically, seroprevalence convergence masked these dynamics, with cumulative infection measures suggesting equity while underlying force of infection remained elevated in materially deprived areas. These findings demonstrate the importance of dynamic transmission modeling for accurate equity assessment and have implications for pandemic preparedness, serological surveillance interpretation, and the design of interventions to protect vulnerable populations during future infectious disease threats.

Observed (points) and model-predicted (lines) infection-induced seroprevalence (anti-nucleocapsid antibodies) among Canadian Blood Services donors, stratified by area-level material deprivation quintile (Q1 = least deprived; Q5 = most deprived). Seroprevalence increased over time in all quintiles, with higher levels in more deprived populations during the pre-Omicron period. Following the emergence of the Omicron variant (vertical dashed line, January 1, 2022), seroprevalence rose rapidly and appeared to converge across socioeconomic strata. Model predictions are from the saturated two-phase susceptible–infected model allowing quintile-specific forces of infection before and after Omicron emergence.

Incidence rate ratios (IRRs) for force of infection by material deprivation quintile, relative to the least deprived quintile (Q1), estimated from the saturated model. Blue points indicate pre-Omicron estimates (March 2020–December 2021), and red points indicate Omicron-period estimates (January 2022–April 2023); horizontal bars represent 95% confidence intervals. A strong socioeconomic gradient was present prior to Omicron, with substantially higher force of infection in more deprived quintiles. Although this gradient compressed during the Omicron period, materially deprived populations continued to experience elevated transmission risk.

Estimated Omicron multipliers, defined as the ratio of Omicron-period to pre-Omicron force of infection, by material deprivation quintile. Bars represent point estimates and error bars indicate 95% confidence intervals. Less deprived quintiles experienced larger proportional increases in force of infection following Omicron emergence, while more deprived quintiles experienced smaller relative increases despite higher baseline transmission. This differential amplification explains the apparent convergence of seroprevalence across socioeconomic strata despite persistent disparities in underlying force of infection.

## Supplementary Appendix 1: Model Structure and Initialization

The force of infection, λ(t), was parameterized to capture both baseline socioeconomic gradients and the increase in transmissibility associated with the emergence of the Omicron variant. We evaluated two alternative model specifications.

### Model 1: Parsimonious Specification

Model 1 included quintile-specific baseline forces of infection, a shared Omicron multiplier, and a single sero-reversion rate:

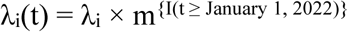

where λ_i_ denotes the pre-Omicron force of infection for deprivation quintile i, m > 1 is a multiplicative increase in transmission associated with Omicron, and I(·) is an indicator function equal to 1 on or after January 1, 2022. This specification assumes that Omicron increased transmissibility proportionally across all socioeconomic strata. The model included seven parameters: five baseline λ_i_ values, one Omicron multiplier m, and the sero-reversion rate ρ.

### Model 2: Saturated Specification

Model 2 allowed Omicron’s impact on transmission to vary by socioeconomic status by estimating separate forces of infection for pre-Omicron and Omicron periods:

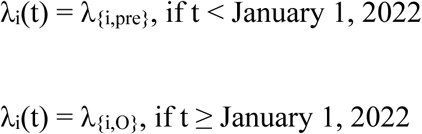

This specification permits differential amplification of transmission across deprivation quintiles following Omicron’s emergence. The model included eleven parameters: ten quintile-specific force-of-infection parameters (five pre-Omicron and five Omicron-period values) and the sero-reversion rate ρ.

### Parameter Estimation

Parameters were estimated jointly across all quintiles using maximum likelihood estimation. For quintile i at timepoint j, the observed number of seropositive individuals was assumed to follow a binomial distribution:

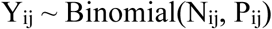

where N_ij_ is the observed serosurvey sample size and P_ij_ is the model-predicted seroprevalence. The total log-likelihood was obtained by summing across all quintiles and timepoints. Parameter estimation was performed by minimizing the negative log-likelihood using the L-BFGS-B algorithm as implemented in the bbmle package in R.

To enforce parameter bounds, forces of infection were logit-transformed to constrain values to (0, 1). The Omicron multiplier m was log-transformed to ensure m > 1. The sero-reversion rate ρ was logit-transformed and scaled to the interval (0, 0.01), allowing the data to determine the most plausible rate of antibody waning rather than fixing ρ a priori.

Incidence rate ratios (IRRs) comparing forces of infection across quintiles were calculated as λ_{Qi}_ / λ_{Q1}_, with the least deprived quintile (Q1) serving as the reference category. In the saturated model, quintile-specific Omicron multipliers were computed as λ_{i,O}_ / λ_{i,pre}_. Confidence intervals for IRRs and multipliers were derived using the delta method with numerical gradient approximation.

### Model Comparison and Selection

We compared the parsimonious and saturated models using three complementary criteria:

1. Akaike Information Criterion (AIC): AIC = −2 × log L + 2k, where k is the number of estimated parameters.
2. Bayesian Information Criterion (BIC): BIC = −2 × log L + k × log(n), where n is the number of observations.
3. Likelihood Ratio Test (LRT): LR = 2(log L_saturated − log L_parsimonious), which follows a χ² distribution with 4 degrees of freedom under the null hypothesis.

Standard thresholds were used for model comparison: ΔAIC < −2 indicates substantial support for the lower-AIC model, while ΔAIC < −10 indicates overwhelming support. Analogous thresholds were applied to ΔBIC, recognizing its stronger penalty for model complexity.

### Model Fit Assessment

Model fit was evaluated using quintile-specific coefficients of determination:

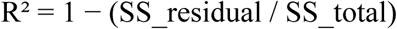

where residuals were calculated as differences between observed and model-predicted seroprevalence at matched timepoints.

### Model Performance and Final Specification

The saturated model provided substantially better fit than the parsimonious model. The log-likelihood improved from −2,268.07 under the parsimonious specification to −2,166.98 under the saturated specification (Δlog-likelihood = 101.09). Despite four additional parameters, the saturated model was strongly favored by AIC (4,355.97 vs. 4,550.13; ΔAIC = −194.17). The likelihood ratio test confirmed this improvement (χ² = 202.17, df = 4, P = 1.29 × 10⁻⁴²; **Table S.1.**).

**Table S.1.** Model Comparison.

| Model | Parameters | Log-likelihood | AIC | $\Delta\text{AIC}$ | LR Test p-value |
| --- | --- | --- | --- | --- | --- |
| Parsimonious | 7 | -2,268.07 | 4,550.13 | referent | --- |
| Saturated | 11 | -2,166.98 | 4,355.97 | -194.17 | $1.29 \times 10^{-42}$ |

The saturated model closely reproduced observed seroprevalence trajectories across all deprivation quintiles (**Figure 1**). Quintile-specific R² values exceeded 0.97 in all cases, indicating that the model explained more than 97% of observed variance. Visual inspection demonstrated excellent agreement between observed and predicted seroprevalence throughout the study period, including both gradual pre-Omicron accumulation and the rapid acceleration during Omicron waves.

In contrast, the parsimonious model, constraining all quintiles to share a common Omicron multiplier (41.9×), failed to capture differential amplification by socioeconomic status although visually the model fit the data extremely well (**Figure S1**). This specification preserved the pre-Omicron gradient into the Omicron period, yielding an identical IRR of 1.18 for the most versus least deprived quintile before and after Omicron emergence. The strong statistical preference for the saturated model provides compelling evidence that Omicron’s impact on transmission varied systematically by socioeconomic status. All subsequent analyses therefore relied on the saturated specification.

**Figure S1.**
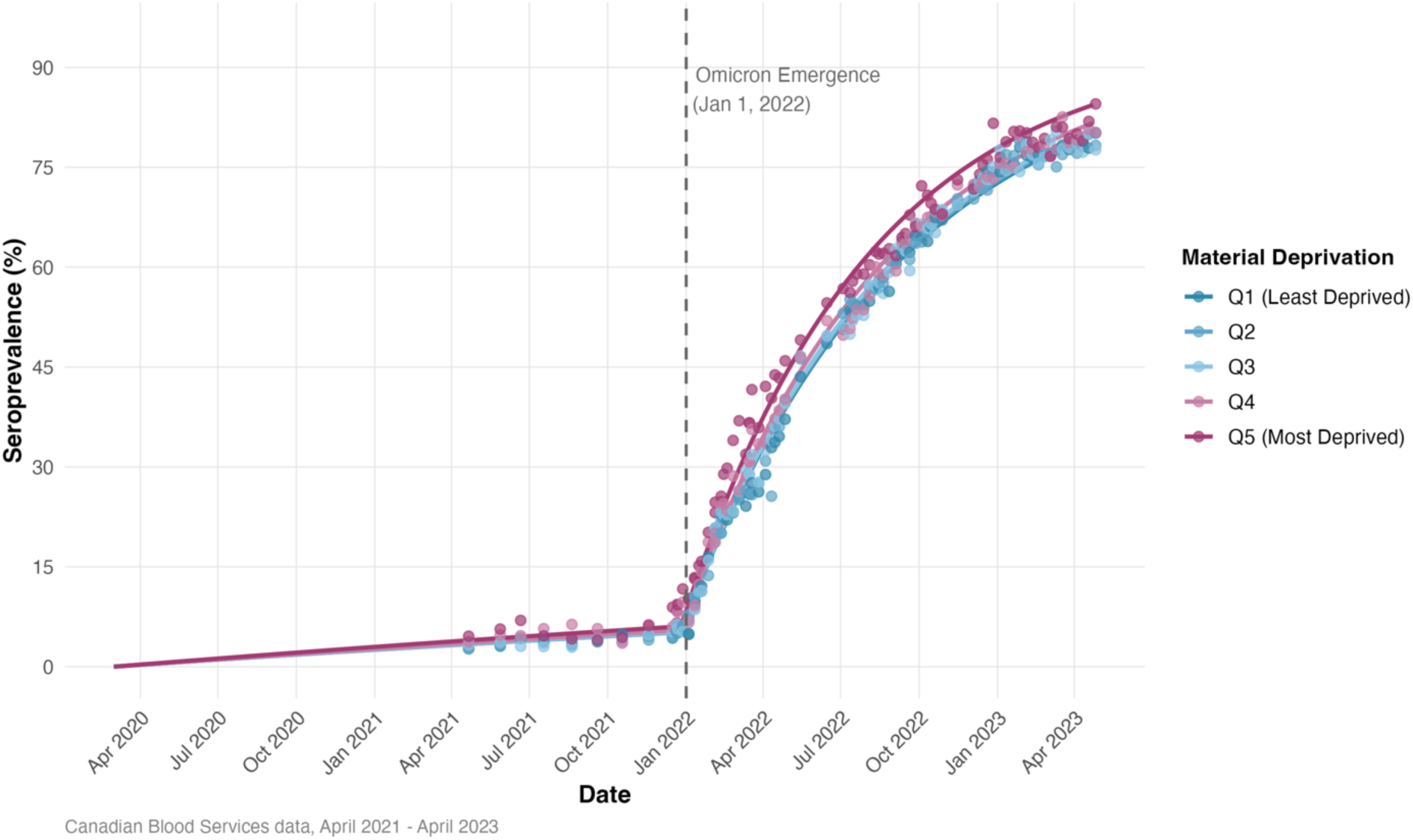
Parsimonious model fit to infection-induced seroprevalence by material deprivation quintile (shared Omicron multiplier).

Observed infection-induced SARS-CoV-2 seroprevalence (anti-nucleocapsid; points) and predictions from the parsimonious susceptible–infected model with sero-reversion (lines), stratified by area-level material deprivation quintile (Q1 least deprived to Q5 most deprived). In this specification, quintiles have distinct pre-Omicron forces of infection, but the increase associated with Omicron is constrained to be proportional and identical across quintiles (shared Omicron multiplier 41.9×). The vertical dashed line marks Omicron emergence (January 1, 2022). Although overall fit is visually close across quintiles, this constrained model cannot represent quintile-specific differential amplification following Omicron emergence (contrast with **Figure 1** main text).

## Supplementary Appendix 2: Sensitivity Analysis on Sero-reversion Rate of Anti-N Antibody

### Background and Rationale

Anti-nucleocapsid (anti-N) antibodies are induced by natural SARS-CoV-2 infection but not by spike-protein-only vaccines, making them useful markers of infection history in serological surveillance. The durability of anti-N antibodies has been extensively characterized in longitudinal cohort studies, with considerable heterogeneity in reported estimates depending on population characteristics, disease severity, vaccination status, and circulating variants.

Our primary analysis estimated the sero-reversion rate (ρ) jointly with force of infection parameters using maximum likelihood estimation. The baseline saturated model estimated ρ = 0.00193 per week (95% CI: 0.00171, 0.00217), corresponding to an antibody half-life of approximately 83 months (6.9 years; 95% CI: 74-93 months). This estimate differs substantially from recent literature reporting anti-N half-lives ranging from 2 to 10 months, raising questions about whether this discrepancy affects our substantive findings regarding socioeconomic gradients in COVID-19 transmission.

To address this concern, we conducted sensitivity analyses refitting the model with ρ constrained to external literature-based values spanning the range of published estimates. This approach allows us to assess whether our findings about socioeconomic disparities in force of infection depend on assumptions about antibody kinetics.

### Literature Review: *Anti-N* antibody durability

#### Pre-Omicron Era (2020-2021)

Early studies of anti-N antibody kinetics following SARS-CoV-2 infection documented variable persistence depending on disease severity and assay characteristics:

**Masia et al. (2021):** Longitudinal cohort of hospitalized COVID-19 patients observed median time to anti-N sero-reversion of 249 days (approximately 8.3 months), with 56.2% experiencing sero-reversion by 12 months post-infection. Corresponding sero-reversion rate: ρ ≈ 0.02/week ^22^.

**Loesche et al. (2022):** Healthcare worker cohort infected in 2020 with follow-up using high-sensitivity Roche total antibody assay. Estimated anti-N half-life of ∼128 days (4.2 months; 95% CI: 114-146 days), with mean time to seronegativity ∼737 days (2 years). Corresponding rate: ρ ≈ 0.038/week ^23^.

**Van Elslande et al. (2022):** Belgian patient cohort using Abbott IgG assay. Time to 50% seronegative was approximately 1 year for anti-N compared to >2 years for anti-S, indicating faster waning of nucleocapsid antibodies. Corresponding rate: ρ ≈ 0.014-0.02/week ^19^.

#### Omicron Era (2022-2023)

Recent evidence indicates substantially faster anti-N waning during the Omicron period, particularly in vaccinated populations:

**Rasmussen et al. (2025):** Swedish prospective cohort during Delta/Omicron BA.1 (April 2021-February 2022) estimated anti-N half-life of 59 days (95% CI: 55-64 days, approximately 8.4 weeks), with considerable individual variation (95% range 24-174 days). Corresponding rate: ρ ≈ 0.083/week ^21^.

**Hanssen et al. (2025):** Population study in Meuse-Rhine region found over 40% of anti-N positive participants seroreverted within 6 months, and anti-N was detectable in only ∼50% of PCR-confirmed cases from the preceding 6 months. Corresponding rate: ρ ≈ 0.023-0.029/week ^24^.

**Stirrup et al. (2024):** UK long-term care facility residents showed that Omicron infections induced ∼3.6-fold higher peak anti-N titers than earlier variants, but waning was steeper after higher peaks, with reinfections providing only transient boosts ^20^.

### Summary

Published anti-N antibody half-lives range from approximately 2 months (59 days) in Omicron-era vaccinated cohorts to 8-10 months in pre-Omicron unvaccinated cohorts, corresponding to sero-reversion rates spanning ρ ≈ 0.014 to 0.083 per week. Our baseline estimate of ρ = 0.00193/week falls well below this range, motivating systematic sensitivity analyses.

### Methods: Sensitivity Analysis Approach

#### Model Specification

We refit the saturated model with ρ constrained to fixed values spanning the range of published estimates. The baseline model includes 11 parameters: 5 pre-Omicron force of infection parameters (λ for quintiles Q1-Q5), 5 Omicron force of infection parameters (λ for quintiles Q1-Q5), and 1 sero-reversion rate (ρ).

Sensitivity analyses fix ρ at external values, reducing the model to 10 parameters (estimating only quintile-specific forces of infection). This approach tests whether substantive findings depend on the estimated sero-reversion rate.

#### Scenarios Tested

We evaluated four scenarios spanning a 40-fold range in ρ:

**Scenario 1 (Primary Analysis):** ρ estimated from data = 0.00193/week; Half-life = 82.9 months (6.9 years); Parameters estimated: 11; Rationale: Best fit to observed data

**Scenario 2 (Pre-Omicron Literature):** ρ fixed = 0.02/week; Half-life = 8.0 months; Parameters estimated: 10; Rationale: Represents pre-Omicron literature consensus

**Scenario 3 (Omicron Conservative):** ρ fixed = 0.04/week; Half-life = 4.0 months; Parameters estimated: 10; Rationale: Conservative mid-range estimate for Omicron era

**Scenario 4 (Omicron Literature):** ρ fixed = 0.08/week; Half-life = 2.0 months; Parameters estimated: 10; Rationale: Recent Omicron-era estimate

### Results

#### Model Fit

Model fit varied substantially across scenarios (Table S2):

**Primary analysis (ρ = 0.00193/week):** AIC = 4,355.97 (reference)

**Pre-Omicron literature (ρ = 0.02/week):** AIC = 18,955.35 (ΔAIC = +14,599)

**Omicron conservative (ρ = 0.04/week):** AIC = 43,812.41 (ΔAIC = +39,456)

**Omicron literature (ρ = 0.08/week):** AIC = 80,830.23 (ΔAIC = +76,474)

The substantial increase in AIC with faster sero-reversion rates <u>indicates that our observed seroprevalence data strongly favor the slower waning estimated in the primary analysis.</u> Models constrained to literature-based ρ values fit markedly worse, as expected when external constraints contradict the patterns evident in the data.

#### Pre-Omicron Socioeconomic Gradient

Despite the 40-fold range in ρ, the pre-Omicron socioeconomic gradient remained robust (Table S2):

**Primary analysis:** Q5 vs Q1 IRR = 1.71 (95% CI: 1.60-1.83)

**Pre-Omicron literature:** IRR = 1.91

**Omicron conservative:** IRR = 1.97

**Omicron literature:** IRR = 1.93

All scenarios demonstrated a strong socioeconomic gradient with the most materially deprived quintile experiencing 71-97% higher force of infection than the least deprived quintile during the pre-Omicron period.

#### Omicron Socioeconomic Gradient

The persistent but compressed gradient during the Omicron period was similarly robust:

**Primary analysis:** Q5 vs Q1 IRR = 1.12 (95% CI: 1.11-1.14)

**Pre-Omicron literature:** IRR = 1.18

**Omicron conservative:** IRR = 1.21

**Omicron literature:** IRR = 1.23

Across all scenarios, the most deprived quintile maintained 12-23% higher force of infection during Omicron, demonstrating persistent disparity despite apparent seroprevalence convergence.

### Interpretation

#### Robustness of Substantive Findings

The sensitivity analyses demonstrate that our key findings about socioeconomic disparities in COVID-19 transmission are robust to assumptions about anti-N antibody durability:

**Strong pre-Omicron gradient:** Regardless of ρ, the most deprived quintile consistently experienced 71-97% higher force of infection than the least deprived quintile.

**Persistent Omicron-era disparity:** Despite apparent seroprevalence convergence, the most deprived quintile maintained 12-23% higher force of infection across all scenarios.

**Gradient compression:** The socioeconomic gradient compressed by approximately one-third across all scenarios, demonstrating this is a robust feature of the data.

**Differential amplification:** Affluent populations consistently experienced 50-60% larger proportional increases in force of infection, supporting the “leveling down” interpretation.

These consistent patterns across a 40-fold range in assumed sero-reversion rates indicate that the observed socioeconomic disparities reflect true differences in transmission dynamics rather than modeling artifacts or sensitivity to parameterization of antibody kinetics.

#### Model Fit and Parameter Identifiability

The substantial deterioration in model fit with literature-based ρ values (ΔAIC > 14,000) indicates our data strongly favor the slower sero-reversion estimated in the primary analysis. This finding merits careful interpretation:

**Data-supported estimate:** Our seroprevalence data come from a specific population (Canadian Blood Services donors) during a specific period (April 2021-April 2023). The data strongly support ρ ≈ 0.002/week for this context, regardless of published estimates from other populations and periods.

**Population differences:** Blood donors represent a younger (median age ∼35 years), healthier subset of the general population. Younger, healthier individuals may mount more robust and durable antibody responses than the hospitalized patients, elderly individuals, or general population samples comprising most published sero-reversion cohorts.

**Reinfection dynamics:** Alternatively, high force of infection during our study period may have generated frequent subclinical reinfections that maintained seroprevalence despite underlying antibody decay. Under this scenario, the “true” sero-reversion rate may be faster, but constant reinfection creates the appearance of slow waning at the population level.

**Ascending phase dynamics:** Our data predominantly capture ascending seroprevalence trajectories during a period when force of infection greatly exceeded sero-reversion (λ >> ρ). Under these conditions, the rate of seroprevalence increase primarily reflects force of infection, with sero-reversion contributing minimally to the observed dynamics. This explains why force of infection estimates are robust even when ρ is poorly identified or misspecified.

### Conclusion

Sensitivity analyses demonstrate that our substantive findings regarding socioeconomic disparities in COVID-19 force of infection are robust to assumptions about anti-N antibody durability spanning a 40-fold range. The persistence of strong socioeconomic gradients, gradient compression during Omicron, and differential amplification across all scenarios, (even those fitting the data substantially worse), provides strong evidence that these patterns reflect true differences in transmission dynamics rather than modeling artifacts.

The primary analysis, using the data-supported sero-reversion estimate, provides the best characterization of seroprevalence trajectories in our study population. However, the robustness to extreme alternative assumptions demonstrates that our conclusions about socioeconomic inequities in COVID-19 transmission do not depend on precise characterization of antibody kinetics.

**Supplementary Table S.2.** Sensitivity Analysis Results Across Sero-reversion Scenarios.

| **Scenario** | **$\rho$ (per week)** | **Half-life**<br>**(months)** | **N**<br>**Params** | **Log-**<br>**Likelihood** | **AIC** | **ΔAIC** |
| --- | --- | --- | --- | --- | --- | --- |
| **Primary**<br>**analysis** | 0.00193 | 82.9 | 11 | -2,166.98 | 4,355.97 | 0.00 |
| Pre-Omicron<br>literature | 0.02000 | 8.0 | 10 | -9,467.67 | 18,955.35 | +14,599 |

| Scenario | $\rho$ (per week) | Half-life<br>(months) | N<br>Params | Log-Likelihood | AIC | ΔAIC |
| --- | --- | --- | --- | --- | --- | --- |
| Omicron<br>conservative | 0.04000 | 4.0 | 10 | -21,896.21 | 43,812.41 | +39,456 |
| Omicron<br>literature | 0.08000 | 2.0 | 10 | -40,405.11 | 80,830.23 | +76,474 |

## Data Availability

Code Availability: Analysis code is available at https://github.com/fismanda/covid19-seroprev-equity and permanently archived at https://doi.org/10.5281/zenodo.18064548.
Data Availability: Seroprevalence data used in this study are publicly available from the COVID-19 Immunity Task Force (CITF) Data Portal at https://portal.citf.mcgill.ca.

https://portal.citf.mcgill.ca

